# High frequency of cerebrospinal fluid autoantibodies in COVID-19 patients with neurological symptoms

**DOI:** 10.1101/2020.07.01.20143214

**Authors:** Christiana Franke, Caroline Ferse, Jakob Kreye, S Momsen Reincke, Elisa Sanchez-Sendin, Andrea Rocco, Mirja Steinbrenner, Stefan Angermair, Sascha Treskatsch, Daniel Zickler, Kai-Uwe Eckardt, Rick Dersch, Jonas Hosp, Heinrich J. Audebert, Matthias Endres, Christoph J. Ploner, Harald Prüss

**Author notes:** **Corresponding Author:** Dr. med. Christiana Franke, Department of Neurology, Charité – Universitätsmedizin Berlin, Campus Benjamin Franklin, Hindenburgdamm 30, 12203 Berlin, Germany.

## Abstract

COVID-19 intensive care patients occasionally develop neurological symptoms. The absence of SARS-CoV-2 in most cerebrospinal fluid (CSF) samples suggests the involvement of further mechanisms including autoimmunity. We therefore determined whether anti-neuronal or anti-glial autoantibodies are present in eleven consecutive severely ill COVID-19 patients presenting with unexplained neurological symptoms. These included myoclonus, cranial nerve involvement, oculomotor disturbance, delirium, dystonia and epileptic seizures. Most patients showed signs of CSF inflammation and increased levels of neurofilament light chain. All patients had anti-neuronal autoantibodies in serum or CSF when assessing a large panel of autoantibodies against intracellular and surface antigens relevant for central nervous system diseases using cell-based assays and indirect immunofluorescence on murine brain sections. Antigens included proteins well-established in clinical routine, such as Yo or NMDA receptor, but also a variety of specific undetermined epitopes on brain sections. These included vessel endothelium, astrocytic proteins and neuropil of basal ganglia, hippocampus or olfactory bulb. The high frequency of autoantibodies targeting the brain in the absence of other explanations suggests a causal relationship to clinical symptoms, in particular to hyperexcitability (myoclonus, seizures). While several underlying autoantigens still await identification in future studies, presence of autoantibodies may explain some aspects of multi-organ disease in COVID-19 and can guide immunotherapy in selected cases.

## Introduction

### Clinical neurological symptoms in COVID-19

A broad variety of neurological symptoms has been observed in COVID-19 patients. Clinical findings comprise hyposmia and hypogeusia in mild cases and agitation, diffuse corticospinal tract signs and myoclonus^1,2^ in severe cases of COVID-19. Neurological syndromes in association with SARS-CoV-2 include many autoimmune diseases, such as Guillain-Barré syndrome (GBS), Miller-Fisher syndrome (MFS), polyneuritis cranialis, meningitis, encephalitis, stroke, epilepsy and myopathy^2,3^. It is debated whether direct virus invasion into the brain can cause pathology, however, SARS-CoV-2 has been detected only scarcely in cerebrospinal fluid (CSF)^3^. Thus, cellular or humoral autoimmunity might contribute to neurological symptoms, similar to other viral diseases. Potential mechanisms include molecular mimicry between viral proteins and neuronal autoantigens and delayed stimulation of post-viral autoimmunity similar to NMDA receptor encephalitis following herpes simplex virus encephalitis (HSE)^4^. We therefore examined the presence of a large panel of anti-neuronal and anti-glial autoantibodies in serum and CSF of COVID-19 patients with predominant neurological symptoms.

## Methods

Between March and May 2020, during the major rise of SARS-CoV-2 infections in Germany, neurological assessment was performed on COVID-19 patients during intensive care unit (ICU) treatment in two tertiary care centers (Charité – Universitätsmedizin Berlin, Campus Virchow-Klinikum (CVK) and Campus Benjamin Franklin (CBF), and Universitätsklinikum Freiburg). In eleven patients with otherwise not explained neurological symptoms lumbar puncture was performed for autoantibody diagnostics in CSF and blood. Written informed consent for research and publication was obtained from all patients or their legal representative (ethics committee approval, Berlin: EA2/066/20, Freiburg: 153/20, laboratory analysis: EA 1/258/18). Autoantibodies against intracellular and surface antigens relevant for central nervous system diseases were measured by line blots, ELISA and cell-based assays (Labor Berlin, Germany) and included antibodies against amphiphysin, CV2 (CRMP5), GAD65, Hu, Ri, Yo, Ma2/Ta, Tr (DNER), GAD65, glutamate receptor (AMPAR1/2, NMDA), DPPX, GABA_A_R, GABA_B_R, mGluR5, LGI1, myelin, Caspr2, dopamine-2 receptor, aquaporin-4, skeletal muscle and phospholipids (cardiolipin, beta2-glycoprotein, annexin). In addition, indirect immunofluorescence on unfixed murine brain sections was performed to search for novel autoantibodies not included in the clinical routine assays, according to established protocols^5^.

## Results

### Patient characteristics

After a median of 12 days [7-17 days] after onset of respiratory symptoms, 11 patients (median age 67 [54-78 years], 8 male) presented with a broad spectrum of neurological symptoms, involving down-beat nystagmus (n=2), other oculomotor disturbances (n=2), aphasia (n=1), hyper- and hypoactive delirium (n=5), partial, mainly orofacial myoclonus (n=6), generalized stimulus-sensitive myoclonus, which improved by sedation and symptomatic treatment (n=1), dystonia of the upper extremities (n=1), stroke (n=1) and epileptic seizures (n=1). Symptoms were not secondary to ICU treatment or explained by infection or internal disease. One patient (#1) had been resuscitated twice for less than two minutes.

### Laboratory findings in SARS-CoV-2 patients with neurological symptoms

CSF and blood samples were analyzed in all patients (Table 1). SARS-CoV-2 PCR in CSF was negative in all patients. Mild pleocytosis was found in 3/11 patients and elevated in one with a positive varicella virus PCR. CSF protein was elevated in 4/11 and matched oligoclonal bands (OCB) present in 6/9 patients.

**Table 1.** Patient characteristics and laboratory findings.

| Patient N° | Sex | Neurological symptoms | SARS-CoV-2 PCR |  | CSF |  |  |  |  | OCB |  | Autoantibody panel<sup>(a)</sup> |  | CSF indirect Immunofluorescence (IgG) |
| --- | --- | --- | --- | --- | --- | --- | --- | --- | --- | --- | --- | --- | --- | --- |
| | | | Swab | CSF | Cell count [$<5/\mu\text{l}$] | Glucose [mg/dl] | Lactate [$<22$ mg/dl] | Protein [$<450$ mg/l] | NfL [pg/ml] | Serum | CSF | Serum | CSF | |
| #1 | m | Downbeat nystagmus, orofacial myoclonus, delirium | pos. | neg. | 8 | 123 | 21 | 594 | $>10,000$ | pos. | pos. | NMDAR IgG 1:1000 | neg. | Medium-sized blood vessels |
| #2 | m | Downbeat nystagmus, generalized stimulus-sensitive myoclonus | pos. | neg. | 1 | 76 | 18,9 | 314 | 5,062 | pos. | pos. | Cardiolipin, beta2 glycoprotein | neg. | Densely clustered pan-neuronal binding |
| #3 | m | Oculomotor disturbance | pos. | neg. | 2 | 113 | 22 | 336 | 1,049 | pos. | pos. | neg. | neg. | Cerebellum granule cells, brainstem neuropil |
| #4 | f | Delirium | pos. | neg. | 5 | 93 | 27,4 | 368 | 1,535 | pos. | pos. | Myelin 1:100 | Neg. | Perinuclear binding |
| #5 | m | Right-sided stimulus-sensitive myoclonus | pos. | neg. | 177 | 83 | 43,8 | 937 | 9,491 | pos. | pos. | neg. | neg. | Cerebellum granule cells, nucleoli of Purkinje neurons |
| #6 | f | Right-sided orofacial myoclonus | pos. | neg. | 17 | 52 | 16,6 | 281 | 2,215 | neg. | neg. | Yo | Yo | Nuclear binding |
| #7 | m | Delirium, myoclonus, epileptic seizures | pos. | neg. | 1 | 75 | 28,1 | 256 | n.d. | n.d. | n.d. | Myelin 1:100 | neg. | Proximal dendrites of Purkinje cells, myelin |
| #8 | m | Right-sided faciobrachial myoclonus, multifocal strokes | pos. | neg. | 1 | 145 | 32,3 | 682 | >10,000 | pos. | pos. | Cardiolipin | n.d. | Blood vessels, glia limitans, astrocytes |
| #9 | m | Oculomotor paresis, transient generalized myoclonus, prolonged awakening | pos. | neg. | 1 | 80 | 16,9 | 741 | n.d. | neg. | neg. | Annexin, cardiolipin | n.d. | Cerebellum granule cells |
| #10 | m | Dystonia right > left upper limb, delirium | pos. | neg. | 1 | 94 | 26,1 | 437,8 | n.d. | n.d. | n.d. | neg. | neg. | Neuropil olfactory bulb, cerebellum, hippocampus |
| #11 | f | Aphasia,<br>neglect,<br>encephalopathy<br>, stupor,<br>impaired<br>consciousness | pos. | neg. | 1 | n.d. | 27 | 168 | n.d. | neg. | neg. | neg. | neg. | Blood<br>vessels,<br>astrocytes,<br>glia<br>limitans |

<sup>(a)</sup> includes antibodies against amphiphysin, CV2 (CRMP5), GAD65, Hu, Ri, Yo, Ma2/Ta, Tr (DNER), GAD65, glutamate receptor (AMPA1/2, NMDA), DPPX, GABA<sub>A</sub>R, GABA<sub>B</sub>R, mGluR5, LGI1, myelin, Caspr2, dopamine-2 receptor, aquaporin-4, skeletal muscle and phospholipids (cardiolipin, beta2-glycoprotein, annexin),
*Abbreviations:* CSF, cerebrospinal fluid; *f*, female; *m*, male; *n.d.*, not determined; *neg*, negative; *OCB*, oligoclonal bands; *pos*, positive.

Using routine diagnostics, one patient showed Yo antibodies in serum and CSF and two patients myelin antibodies in serum. One patient had high-level serum IgG NMDA receptor antibodies. Neurofilament light chain (NfL) levels in CSF were increased in all tested patients (7/7).

### Screening assay for novel CSF autoantibodies in SARS-Cov-2 positive patients

CSF analysis for the presence of anti-neuronal autoantibodies not included in commercial routine assays using indirect immunofluorescence on unfixed mouse brain sections reproducibly showed strong IgG binding in most patients. IgG staining patterns included vessel endothelium, perinuclear antigens, astrocytic proteins and neuropil of basal ganglia, hippocampus or olfactory bulb (Fig. 1). Although antigenic epitopes are currently unknown, the intense staining indicates high specificity to certain neuronal, astrocytic and vascular proteins.

**Figure 1.**
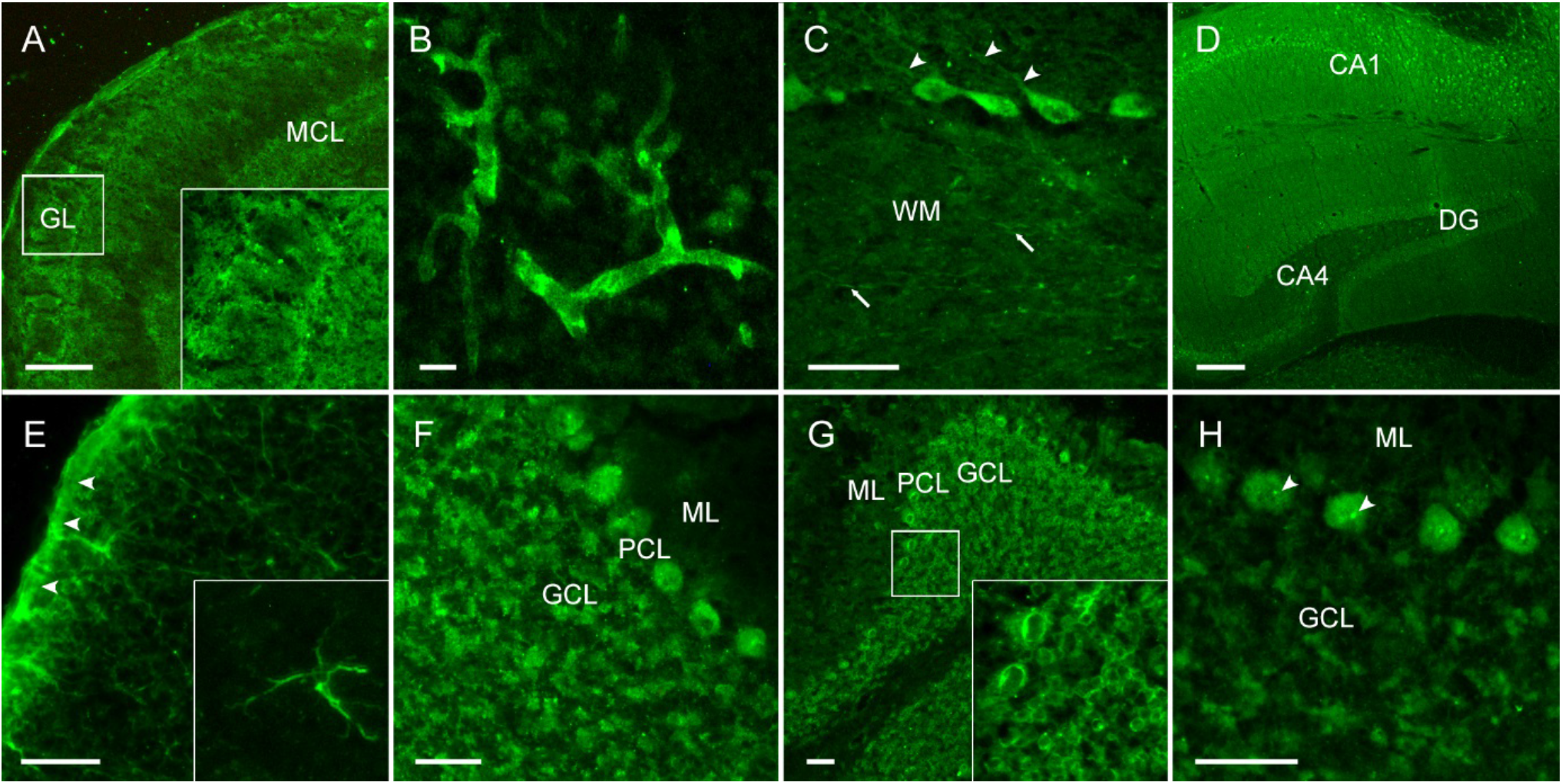
CSF of COVID-19 patients shows strong IgG autoreactivity on unfixed mouse brain sections. Representative images of indirect immunofluorescence demonstrate autoantibody binding to circumscribed anatomical structures including (A) neuropil of the olfactory bulb, (B) medium-sized vessels in the brain, (C) proximal dendrites of Purkinje neurons (arrowheads) and myelinated fibers (arrows) in the cerebellum, (D) neuropil in the hippocampus, (E) glia limitans (arrowheads) and astrocytes (enlarged box) throughout the brain. Several autoantibodies target intracellular antigens, such as (F) densely clustered intraneuronal epitopes, (G) perinuclear antigens or (H) nucleoli (arrowheads) as part of an anti-nuclear antibody response. *Abbreviations: CA1/4*, cornu ammonis 1/4; *DG*, dentate gyrus; *GCL*, granule cell layer; *GL*, glomerular layer; *MCL*, mitral cell layer; *ML*, molecular layer; *PCL*, Purkinje cell layer; *WM*, white matter. Scale bars: 100 µm (A, E), 50 µm (B-C, F-H) and 250 µm (D).

## Discussion

We report autoantibody findings in eleven critically ill COVID-19 patients presenting with a variety of neurological symptoms with unexplained etiology. Cardiopulmonary resuscitation was required after asystole in one patient with high-level serum IgG autoantibodies against NMDA receptors, possibly reflecting NMDA receptor encephalitis, in which arrhythmia and autonomic dysfunctions are common. Symptomatic treatment with valproic acid and clonazepam was successfully administered in two patients with myoclonus. Due to the retrospective nature of this study, our findings could not guide treating physicians to initiate immunomodulatory therapy. Recently, clinical improvement of COVID-19 patients with GBS has been reported after therapy with intravenous immunoglobulins (ivIG)^6^ and after steroids in COVID-19 patients with encephalitis^7^, indicating that immunotherapy should be considered in future cases of CSF autoantibody-positive COVID-19 patients. In most patients, increased CSF protein, lactate or white blood cells with negative SARS-CoV-2 PCR indicated inflammatory changes compatible with autoimmune encephalitis. NfL was markedly elevated (>5,000 pg/ml) in CSF of four COVID-19 patients, thus exceeding established cut-off values (>2,500 pg/ml) for rapidly progressing neurodegenerative diseases such as amyotrophic lateral sclerosis and multisystem atrophy^8^, and also found in autoimmune encephalitis^9^. Elevated NfL levels might reflect direct tissue destruction from viral replication or from inflammatory damage. Whether this is a transient elevation or a continuous transformation into a degenerative phenotype is yet to be determined^10^. The high frequency of CSF anti-neuronal and anti-glial autoantibodies is remarkable, as is the confinement to specific immunofluorescence patterns (Fig. 1). Although more than one patient each had IgG autoantibodies targeting neuropil, astrocytes or medium-sized blood vessels, it will require larger patient cohorts for linking a given autoantibody pattern to clinical symptoms. We regularly encounter similar immunofluorescence patterns in patients with autoimmune encephalitis, but not in healthy control CSF. As most of these novel autoantigens are yet to be determined, it is challenging to judge whether CSF autoantibodies in COVID-19 are pathogenic or not. The neuropil pattern in some patients suggests binding to surface receptors or ion channels and thus pathogenicity (Fig. 1A, D), similar to the rapidly growing group of antibody-mediated encephalitides^11^. Likewise, the astrocyte pattern in two patients (Fig. 1E) is reminiscent of the relatively common form of GFAP antibody encephalitis^12^. Other antigens, however, are located intracellularly (Fig. 1F-H), indicating that the humoral immune response is secondary to other immune mechanisms, including CNS damage from cytotoxic T cells and innate autoimmunity.

Post-viral humoral autoimmunity is an emerging concept best studied for NMDA receptor encephalitis developing in almost 30% of cases post HSE. Tissue destruction may lead to the release of brain-restricted ‘neo-antigens’ such as NMDA receptors, and viral material might provide costimulatory signals to antibody-producing cells^13^. Recent findings suggest that several viral infections can lead to secondary autoimmune encephalitis, including EBV, HHV-6, enterovirus, adenovirus, hepatitis C or HIV infections^14^. Thus, the present findings suggest that SARS-CoV-2 is no exception to this general principle. The following months will show whether such autoreactivity can cause persisting neurological morbidity even after clearance of SARS-CoV-2 and remission of COVID-19, in a way reminiscent of the unexplained severe ‘encephalitis lethargica’ – commonly with postencephalitic parkinsonism – in more than a million patients of the influenza pandemic in 1918^15^. Together, the high frequency of autoantibodies targeting the brain in the absence of other explanations suggests a causal association with clinical symptoms, in particular with hyperexcitability (myoclonus, seizures). While several underlying autoantigens still await identification in future studies, presence of autoantibodies may explain some aspects of multi-organ disease in COVID-19 and guide immunotherapy in selected cases.

## Data Availability

All data is available by request.

